# Characterization of highly active mutational signatures in tumors from a large Chinese population

**DOI:** 10.1101/2023.11.03.23297964

**Authors:** Aaron Chevalier, Tao Guo, Natasha Q. Gurevich, Jingwen Xu, Masanao Yajima, Joshua D. Campbell

## Abstract

The majority of mutational signatures have been characterized in tumors from Western countries and the degree to which mutational signatures are similar or different in Eastern populations has not been fully explored. We leveraged a large-scale clinical sequencing cohort of tumors from a Chinese population containing 25 tumor types and found that the highly active mutational signatures were similar to those previously characterized^1,2^. The aristolochic acid signature SBS22 was observed in four soft tissue sarcomas and the POLE-associated signature SBS10 was observed in a gallbladder carcinoma. In lung adenocarcinoma, the polycyclic aromatic hydrocarbon (PAH) signature SBS4 was significantly higher in males compared to females but not associated with smoking status. The UV-associated signature SBS7 was significantly lower in cutaneous melanomas from the Chinese population compared to a similar American cohort. Overall, these results add to our understanding of the mutational processes that contribute to tumors from the Chinese population.

## Main

A variety of exogenous exposures or endogenous biological processes can contribute to the overall mutational load observed in human tumors^1,3,4^. Many different mutational patterns, or “mutational signatures”, have been identified across different tumor types^3,5–9^ which can provide a record of environmental exposure or clues about the etiology of carcinogenesis^10^. The majority of mutational signature characterization has been performed using tumors from Western populations due to the availability of sequencing data in these regions from large-scale atlases such as The Cancer Genome Atlas (TCGA)^11^ and the International Cancer Genome Consortium (ICGC)^12^. While cancer incidence and mortality can vary across regions and countries^13^, the degree to which mutational signatures are similar or different between populations remains an open area of exploration. Exposures to certain carcinogens and their corresponding mutational signatures have been characterized in Eastern populations. For example, aristolochic acid is found in herbal medicines used in Asian populations and can cause the single-base substitution (SBS) signature SBS22 in hepatocellular carcinomas (HCCs)^14^. Similarly, exposure to aflatoxin B1 produced by molds growing on food can cause SBS24 in HCCs^15^.

Previously, a large-scale cohort of tumors from the Chinese population was profiled with a targeted DNA sequencing panel developed by OrigiMed (OM cohort)^16^. This work compared the frequency of driver genes, tumor mutational burden (TMB), gene fusions, and clinically actionable alterations between tumors from Chinese and American populations. We further leveraged this cohort to characterize the landscape of highly active mutational signatures across a large spectrum of tumor types in the Chinese population. 2,115 tumors with at least 10 mutations from 25 tumor types were utilized for mutational signature analysis. We first performed *de novo* deconvolution with Non-Negative Matrix Factorization (NMF) using the SigProfiler package^17^ and identified six mutational signatures (**Supplementary Figure 1, Supplementary Table 1, Supplementary Table 2**). All signatures were highly correlated with previously defined signatures in the COSMIC database, indicating that no new highly active mutational processes were present in this cohort. The discovered signatures included those correlated with SBS1/6, SBS4, SBS10, SBS12/26, SBS2/13, and SBS22.

The limited number of mutations in the targeted sequencing data can hinder *de novo* mutational signature discovery. Therefore, we also predicted signature activity levels for existing COSMIC signatures using the musicatk package^18^ (**Supplementary Table 3**). The landscape of activities for 16 signatures across tumor types is shown in **Figure 1** (**Supplementary Table 4**). Signatures related to endogenous biological processes included the aging-related signature SBS1 and the clock-like signature SBS5 which were broadly detected across tumor types. The APOBEC-related signatures SBS2 and SBS13 were often observed together across a variety of epithelial tumor types such as breast carcinoma, carcinoma of the uterine cervix, esophageal carcinoma, gall bladder carcinoma, non-small cell lung cancer (NSCLC), and urothelial carcinoma. Several signatures related to defective mismatch repair (MMR), microsatellite instability (MSI), or defective DNA polymerase activity were detected, including SBS6, SBS15, SBS20, SBS21, SBS26, SBS17, and SBS10. As previously observed in Western cohorts, these signatures often co-occurred in the same samples^1,2^. One or more of these signatures were active in samples from many of the same tumor types observed in Western cohorts including bone sarcoma^1^, colorectal carcinoma^1^, intrahepatic and extrahepatic cholangiocarcinoma^1^, gall bladder carcinoma^19^, gastric cancer^20^, head and neck carcinoma^1^, kidney/renal cell carcinoma^1^, ovarian carcinoma^1^, pancreatic cancer^1^, small bowel carcinoma^21^, soft tissue sarcoma^22^, and uterine corpus endometrial carcinoma^1^. We also observed three gallbladder carcinomas with high levels of SBS6 as well as one sample with SBS10 (POLE), which has not been previously observed for this cancer type^23,24^. High levels of SBS17 were found in a small number of tumors from breast carcinoma, colorectal carcinoma, extrahepatic cholangiocarcinoma, gastric cancer, or liver/hepatocellular carcinoma. While this signature often co-occurred with other signatures, some tumors contained only this signature. High levels of SBS10 related to defects in POLE were observed in 22 colorectal carcinomas and 3 uterine corpus endometrial carcinomas.

**Figure 1.**
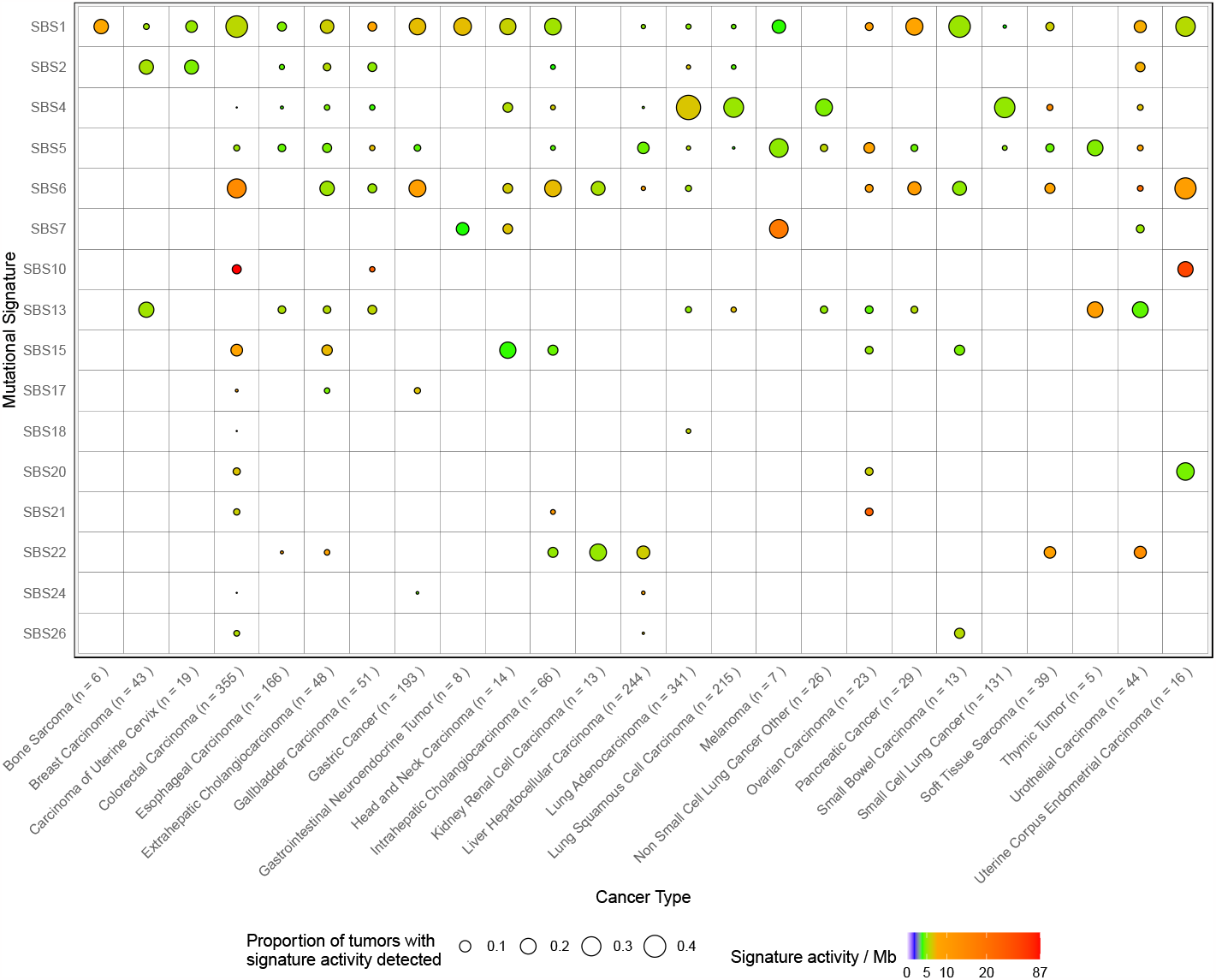
The landscape of highly active mutational signatures in a large Chinese population. Sixteen mutational signatures were identified across 2,115 tumors from 25 tumor types. The size of the dot corresponds to the percentage of tumors within each tumor type that have detectable levels of the signature. Signatures with at least 10 counts were considered detected in an individual tumor. The color of the dot corresponds to the median activity of each signature within the detected samples per megabase (Mb).

Several detected signatures are known to be caused by exposure to exogenous DNA damaging agents. SBS4 is caused by exposure to polycyclic aromatic hydrocarbons (PAHs) such as benzo[a]pyrene in cigarette smoke^25^ and was observed in small cell lung cancers (SCLCs) and NSCLCs. SBS22 is caused by exposure to aristolochic acid and was detected in liver/hepatocellular carcinomas, intra and extrahepatic cholangiocarcinomas, kidney/renal cell carcinomas, and urothelial carcinomas (**Figure 2A**). Interestingly, SBS22 was also detected in four soft tissue sarcomas and one esophageal carcinoma, which has not been previously reported ^22,26^ (**Figure 2B**). SBS24 reflecting aflatoxin B1 exposure was detected at high or moderate levels in 2 liver/hepatocellular carcinomas similar to previous reports^1,27^. This signature was also highly detected in 1 colorectal carcinoma and 1 gastric cancer raising the possibility that this exposure or something similar can induce mutations in more tumor types than previously appreciated. SBS7 is caused by UV radiation exposure and was highly detected in 2 melanomas, 1 head & neck carcinoma, and 2 urothelial carcinomas. Lastly, SBS18 which is potentially due to oxidative stress was detected in 1 colorectal carcinoma.

**Figure 2.**
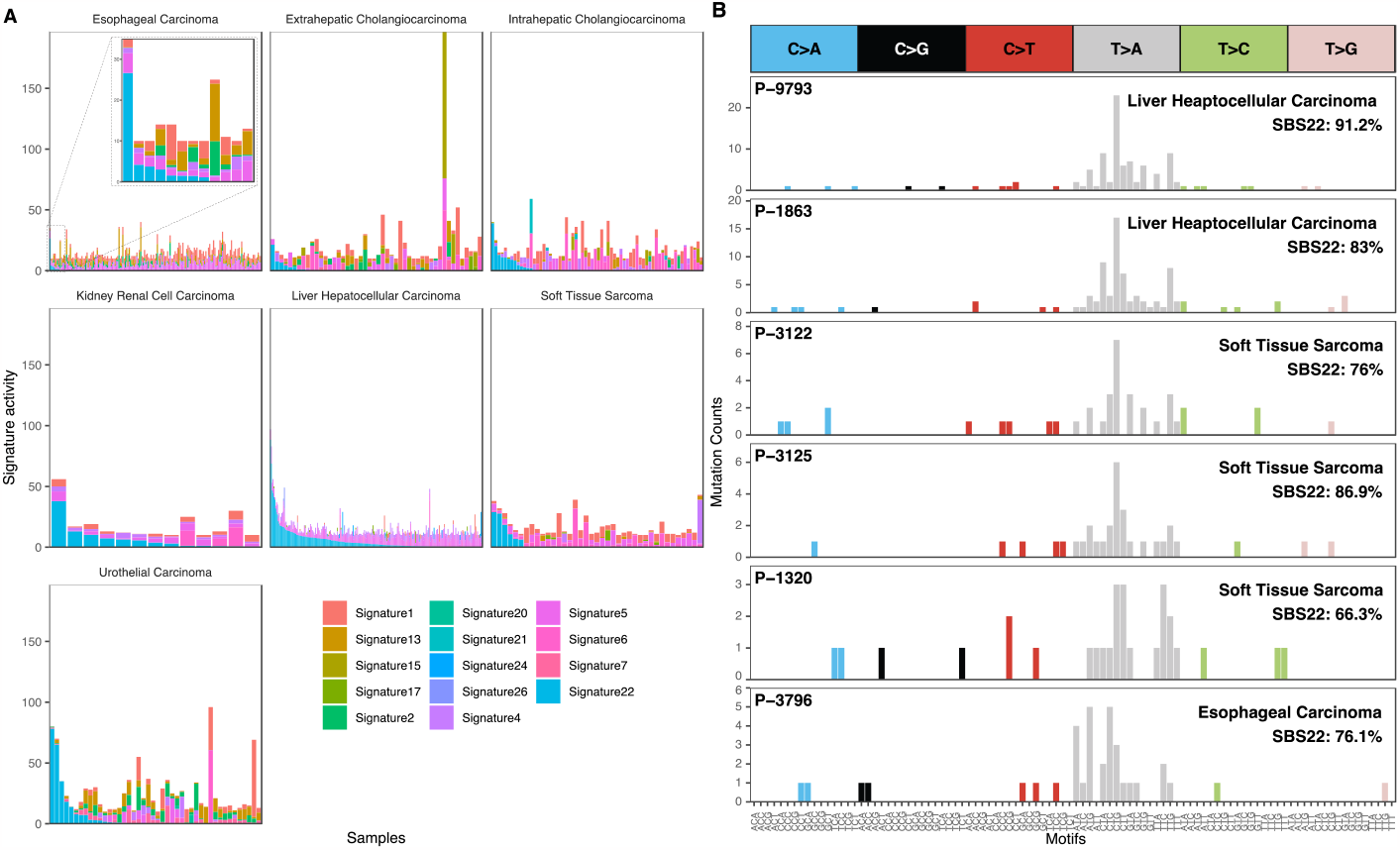
Prevalence of aristolochic acid (SBS22) activity across tumor types. **(A)** SBS22 reflects exposure to aristolochic acid and was highly active in 32 liver/hepatocellular carcinomas, 6 intra or extrahepatic cholangiocarcinomas, 3 kidney renal cell carcinomas, and 5 urothelial carcinomas. This signature was also detected in 4 soft tissue sarcomas and 1 esophageal carcinoma which has not been reported in Western cohorts. Each bar represents the level of the estimated signature activities in each sample and samples are ordered by the level of SBS22. **(B)** Mutation counts for 2 example liver/hepatocellular carcinomas, 3 soft tissue sarcomas, and 1 esophageal carcinoma that predominantly contain the SBS22 signature.

We next sought to directly compare the signature activity levels in this Chinese population to a similar clinical cohort of tumors from an American population. Specifically, we leveraged a dataset generated at Memorial Sloan Kettering (MSK) that was profiled with the IMPACT targeted sequencing panel. After mapping major tumor types and tumor subtypes between cohorts, 13 groups with at least 5 samples in each cohort were compared (**Supplementary Table 5**). For each signature, we first compared the proportion of samples with high activity of that signature between cohorts using a Fisher’s exact text. The only significant difference was that the American cohort had a higher proportion of tumors with the UV signature SBS7 in soft tissue sarcoma compared to the Chinese cohort (FDR < 0.05; **Figure 3A; Supplementary Table 6**). The MSK cohort did not have any tumors with high SBS22 reflecting that aristolochic acid exposure is largely occurs in Eastern population. Next, we compared the median activity of the proposal signature activities across cohorts. The majority of the proportional signature activities were not significantly different between cohorts after applying an FDR correction (**Figure 3B**; **Supplementary Table 7**). The only exceptions were that SBS2 in breast carcinoma, SBS6 in colorectal carcinoma, and SBS5 in hepatocellular carcinoma had significantly lower levels in the OM cohort compared to the MSK cohort (Wilcoxon rank-sum test; FDR < 0.05). These results demonstrate that the major differences between populations are due to exposure to specific mutagens and that the levels of other common signatures are largely the same in tumors across Chinese and American populations despite the underlying heterogeneity in clinical and regional characteristics.

**Figure 3.**
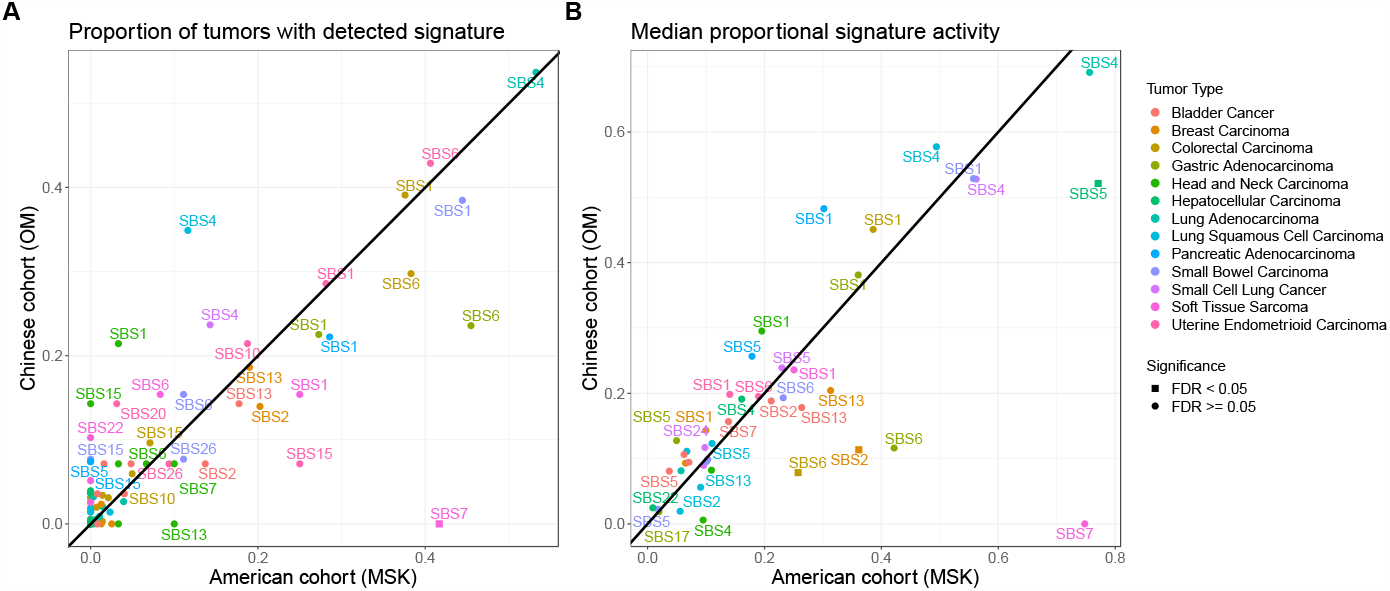
Comparison of signature detection and activity levels across populations. **(A)** Signatures with at least 10 counts in a tumor were considered detected in that tumor. The frequencies of detected signatures were compared across Chinese (OM) and American (MSK) cohort using the Fisher’s exact test with an FDR correction. Only tumor types with at least 5 tumors in each cohort were included. The majority of signature detection rates were not significantly different between cohorts with the exception of the UV-associated signature SBS7 which was found in a higher proportion of soft tissue sarcomas in the MSK cohort compared to the OM cohort. **(B)** The median levels of proportional signature activities were compared for matched tumor types between the Chinese (OM) and American (MSK) cohorts using a Wilcoxon rank-sum test with an FDR correction. The majority of signature activities were not significantly different between cohorts with the exception of the APOBEC SBS2 signature in breast carcinoma, the SBS6 signature in colorectal carcinoma, and the SBS5 signature in hepatocellular carcinoma.

SBS4 was highly prevalent across NSCLCs including lung adenocarcinomas (LUADs) and lung squamous cell carcinomas (LUSCs) as well as in small cell lung cancer (SCLCs) as previously observed^1,25,28,29^ (**Figure 1, Figure 4A**). In China, rates of cigarette smoking are much higher among males compared to females^30^. We observed similar trends in this cohort with a higher proportion of male in the smokers compared to the non-smokers (p < 2.2e-16; Fisher’s exact text; **Figure 4B**). Furthermore, LUADs from female non-smokers in Asian populations tend to be driven by alterations in *EGFR*^31^. Similarly, in this cohort we observed a higher proportion of tumors with *EGFR* mutations in females compared to males in both the smokers (p = 0.0061) as well as the non-smokers (p = 0.0004; **Figure 4B**). In Western cohorts, SBS4 activity is strongly associated with smoking status in LUAD^32^. Surprisingly in this Chinese cohort, the SBS4 signature was not associated with smoking status but instead was strongly associated with sex (**Figure 4C**). Specifically, the level of SBS4 activity was significantly higher in males compared to females within smokers (p = 0.00398) and even more so within non-smokers (p = 4.37e-07). SBS4 was not significantly different between smokers and non-smokers in males or between smokers and non-smokers in females (p > 0.05) indicating that the association with sex is not just due to the differences in the prevalence of smoking between men and women. When applying a multivariate linear model to a subset of LUADs with complete clinical information (n=271), SBS4 activity was significantly higher in males compared to females (p = 1.48e-07), decreasing with age (p = 0. 0.0013), lower in tumors with *EGFR* mutations (p = 0.0072), and lower in metastatic versus primary samples (p = 0.0004). SBS4 activity was not significantly associated with tumor purity, stage, or smoking status (p > 0.01; **Figure 4D**). No associations were found between SBS4 activity and these variables in LUSC or SCLC (p > 0.01; **Supplementary Figure 2**) demonstrating that this phenomenon is specific to LUAD. While inaccurate self-reported smoking status may be a factor, these findings may suggest that factors other than smoking status are contributing to SBS4 mutations in males from China. Other possible factors affecting SBS4 mutations in males may include a higher daily usage of cigarettes or a higher exposure to air pollution and ambient particulate matter compared to women ^33,34^. Additional cohorts of LUADs with detailed smoking and exposure history will likely be needed to further understand the causes of this association.

**Figure 4.**
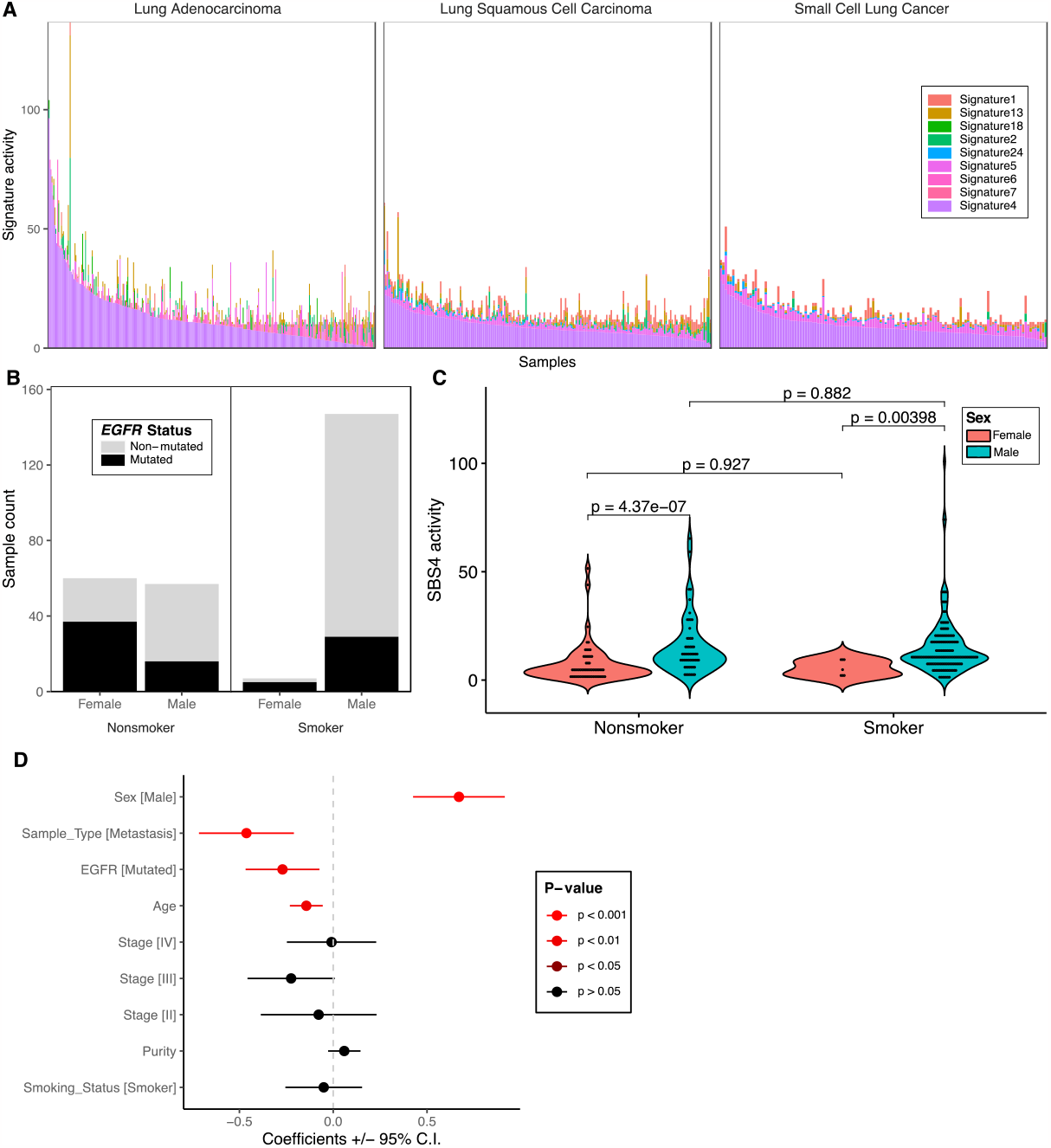
SBS4 activity associated with sex but not smoking status in lung adenocarcinoma. **(A)** SBS4 activity was predominantly observed in lung cancers including lung adenocarcinoma (LUAD), lung squamous cell carcinoma (LUSC), and small cell lung cancer (SCLC). Each stacked bar represents the estimated activity levels of each signature in each tumor. Signatures with no estimated activity in these tumor types were excluded. **(B)** The relationship between smoking status, sex, and *EGFR* mutation status is shown for a subset of LUADs with complete clinical information (n=271). A higher proportion of males were observed in smokers compared to non-smokers and a higher proportion of *EGFR* mutations were observed in females compared to males in both smokers and non-smokers. **(C)** Using a Wilcoxon rank-sum test, SBS4 activity was significantly higher in males compared to females in both smokers and non-smokers but was not significantly different between smokers and nonsmokers within males or within females. **(D)** Using a multivariate linear model, SBS4 activity was associated with sex, sample type (i.e. lower in metastasis compared to primary), *EGFR* mutation status, and age.

In the OM cohort, only 7 of 54 melanomas had greater than 10 mutations (13%) which is significantly less than the MSK cohort in which 145 of 358 (41%) of the melanomas had greater than 10 mutations (p = 6.0e-05; Fisher’s exact text). SBS7 was only highly detected in 2 of the 7 melanomas in the OM cohort (**Figure 5A**). Asian populations have higher rates of acral and mucosal melanomas whereas Western populations have higher rates of cutaneous melanoma^35,36^. Acral and mucosal subtypes often have lower mutation rates and are less driven by UV-induced DNA damage compared to cutaneous melanomas^37^. To understand differences between populations specifically in cutaneous melanoma, we expanded the analysis to examine the mutational profiles of all cutaneous melanomas across both cohorts (including tumors with less than 10 mutations which were excluded in the mutational signature analysis). Cutaneous melanomas from the OM cohort (n=26) had significantly lower SBS mutations per megabase (Mb) than cutaneous melanomas from the MSK cohort (n = 191; p = 1.5e-13; **Figure 5B**). They also had lower frequencies of any C>T mutations (p = 1.1e-7; **Figure 5C**) and C>T mutations at the TCA, CCC, and TCT trinucleotide contexts, which are the most common contexts in the UV-associated signature SBS7 (p = 1.1e-5; **Figure 5D**). These findings corroborate the recent work showing that the number of UV-associated mutations in normal skin is lower in Asian populations compared to Western populations despite Asian populations having higher levels of exposure to UV radiation ^38^. Additionally, lower response rates to immune checkpoint inhibitors have been observed in melanomas from the Chinese population compared to Western populations^39^ which could be a result of the lower tumor mutation burden (TMB)^40^. Our data suggests that the lower prevalence of UV-associated mutations is a major contributor to the lower overall TMB in Chinese cutaneous melanomas.

**Figure 5.**
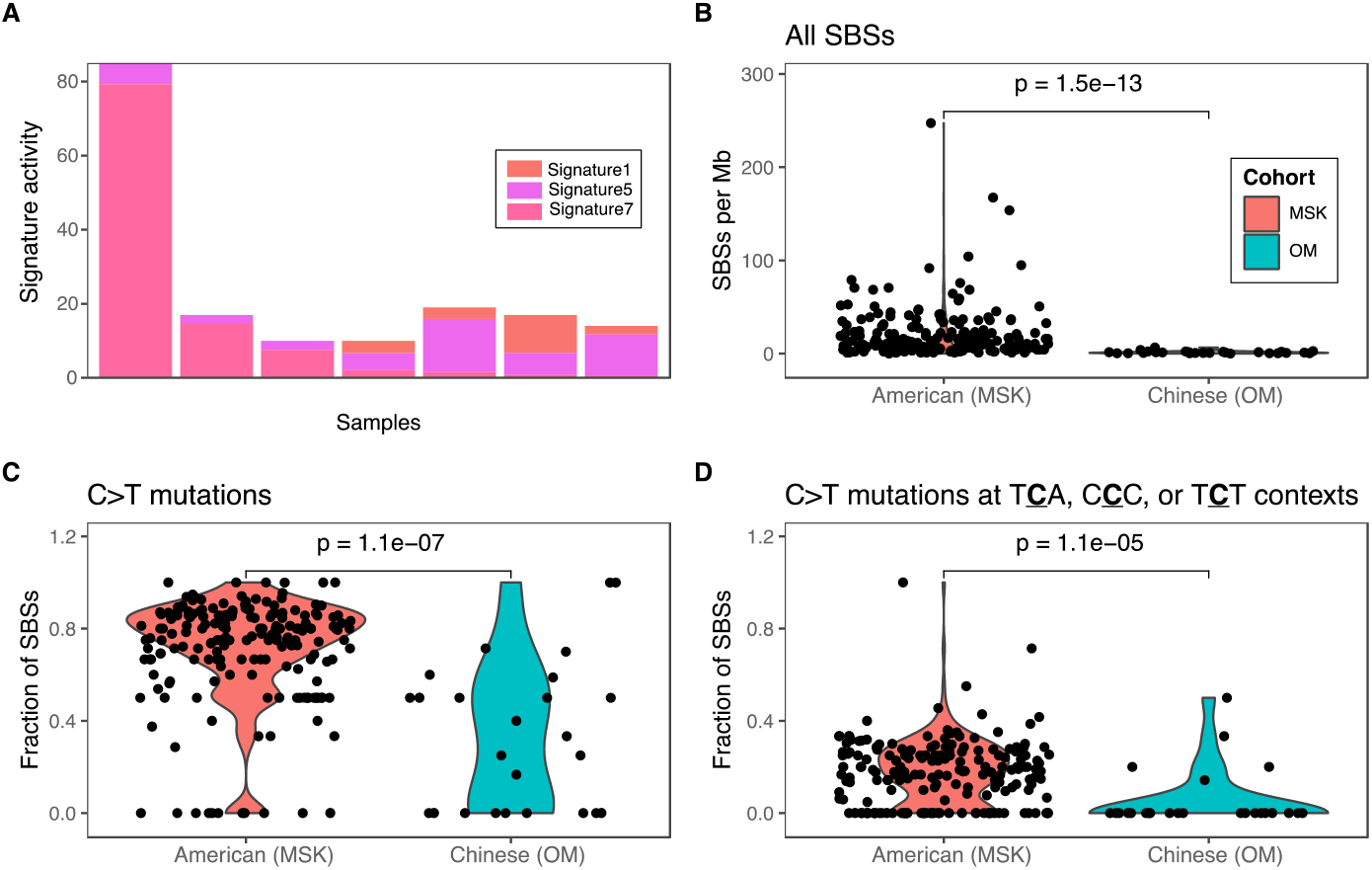
Decrease in UV-associated mutations in cutaneous melanomas from Chinese patients. **(A)** Barplot of signature activities for 7 melanomas in the OM cohort. **(B)** The total number of SBS mutations per megabase (Mb), **(C)** the fraction of C>T mutations, and **(D)** the fraction of C>T mutations at the T<u>**C**</u>A, C<u>**C**</u>C, or T<u>**C**</u>T trinucleotide contexts were significantly lower in cutaneous melanomas from the Chinese cohort compared to cutaneous melanomas from the American cohort.

Overall, this analysis provides an overview into the mutational signatures that are highly active in a large Chinese population. One limitation of this study is that the OM cohort was profiled with a targeted sequencing panel which has a lower number of mutations detected per tumor. Having lower counts can hinder detection of signatures that tend to have lower activity levels. For example, we were not able to confidently detect signature SBS3 which denotes homologous repair deficiency (HRD) caused by loss of *BRCA1/2*. SBS3 has been previously characterized in breast cancers from Chinese and Korean populations^41,42^. Despite this limitation, we were able to detect signatures that are highly active in these tumors (i.e. signatures that produce enough mutations to be detected with a limited targeted sequencing panel) and characterize novel associations specific to this Chinese population.

## Online Methods

### Cohort

The full details of the patient consent, clinical characteristics, biospecimen processing, and DNA sequencing have been previously described^16^. Briefly, tumors from 25 tumor types were profiled with a DNA sequencing panel targeting 450 genes, *TERT* promoter mutations, and 39 introns in a Clinical Laboratory Improvement Amendments (CLIA)-certified and College of American Pathologists (CAP)-accredited laboratory by the Chinese-based company OrigiMed. The mutations and clinical data were retrieved from the cBioPortal (https://www.cbioportal.org/study/summary?id=pan_origimed_2020). Each tumor has a major cancer type and a more specific detailed cancer type. For most tumors, we used the major cancer type label. Given the high numbers of Non-Small Cell Lung Cancers (NSCLCs) in this dataset, we divided this group into Lung Adenocarcinomas (LUADs), Lung Squamous Cell Carcinomas (LUSCs), and “NSCLC-Other” which contained the subcategories of “Lung Adenosquamous Carcinoma”, “Large Cell Lung Carcinoma”, and “Non-Small Cell Lung Cancer Other”.

### Signature discovery and prediction

Single base substitutions (SBSs) were extracted in each tumor to produce a count table using the musicatk R package v1.91^18^. Mutational signature discovery and prediction was limited to 2,115 tumors from 25 major tumor types that had at least 10 single base substitutions. Mutational signatures were first discovered *de novo* using NMF from the SigProfiler package v1.1.4^17^ setting the reference genome to GRch38. The number of signatures predicted was varied from one to eight and the optimal number of six was chosen based on the maximal difference between the mean sample cosine distance and average stability metrics. Discovered signatures were compared to those from the COSMIC V2 database using cosine similarity. Next, the activity of all COSMIC V2 signatures were predicted using the “auto_predict_grid” function in the musicatk package with the “algorithm” parameter set to “lda_posterior” and the “sample_annotation” parameter set to the tumor type^18^. Initially, 25 signatures were detected with this method. We manually reviewed each signature according to criteria set in the PanCancer Analysis of Whole-Genomes (PCAWG) mutational signature working group^2^. Specifically, we examined the mutation counts from individual tumors with high predicted activities of each signature. If no individual tumors displayed a mutational pattern that was predominantly correlated with the signature, then this signature was excluded from the final signature set. The final signature activities were predicted using the “predict_exposure” function with the “algorithm” parameter set to “lda” and specifying sixteen SBS signatures that passed inspection including 1, 2, 4, 5, 6, 7, 10, 13, 15, 17, 18, 20, 21, 22, 24, and 26. A signature was considered highly active in an individual tumor if it contained at least 10 estimated counts in that tumor. Proportional signature activities were calculated by dividing each signature activity within a tumor by the total number of estimated signature activity counts from that tumor.

### Comparison to Western cohorts

Mutations for the MSK cohort were downloaded from cBioPortal (https://www.cbioportal.org/study/summary?id=msk_impact_2017). SBSs were extracted from each tumor using the musicatk package and samples with at least 10 mutations were retained for the analysis. The same signatures that were detected in the OM cohort were predicted in the MSK cohort using the “predict_exposure” function with the “algorithm” parameter set to “lda”. Tumor types were mapped between cohorts using either the major or more detailed cancer type label (**Supplementary Table 3**). The medial level of proportional activity for each signature in a tumor type was compared across cohorts using a Wilcoxon rank-sum test followed by correction for multiple hypothesis testing using the False Discovery Rate (FDR). The frequencies of tumors with detected signature activity were compared across cohorts using the Fisher’s exact test followed by and FDR correction. Only tumor types with at least 5 tumors in both cohorts were including in the analysis. For both comparisons, only signatures with a median level of 0.01 across all samples from both cohorts were included in the analysis.

### Determining association to other clinical or genomic variables

The Wilcoxon rank-sum test was used to assess differences in normalized signature activities between two groups. The association between SBS4 activity and covariates was also assessed using multivariate linear regression within each lung tumor type. SBS4 activity was log transformed after adding a pseudocount of 1 and treated as the dependent variable. Independent variables included age at diagnosis, sex, smoking status, stage according to the American Joint Committee on Cancer (AJCC), tumor purity, *EGFR* mutation status, and sample type (primary or metastasis). Samples with “Unknown” status for any variable were excluded from the regression and any variable that did not have more than one category within a cancer type was excluded from the regression for that cancer type.

## Data Availability

The data were openly available before publication of our study, from OrigiMed and MSK and are respectively available at the following links:
https://www.cbioportal.org/study/summary?id=pan_origimed_2020
https://www.cbioportal.org/study/summary?id=msk_impact_2017

## Code availability

All code for the analysis is available on GitHub at https://github.com/campbio-manuscripts/Chinese_Mutsigs.

## Acknowledgements

This work was funded by the National Cancer Institute (NCI) Informatics Technology for Cancer Research (ITCR) 1U01CA253500 (J.D. Campbell and M. Yajima). We thank Drs. Kai Wang and Xiaoliang Shi for helpful insights about the OrigiMed dataset.

## Supplementary Figures

**Supplementary Figure 1.**
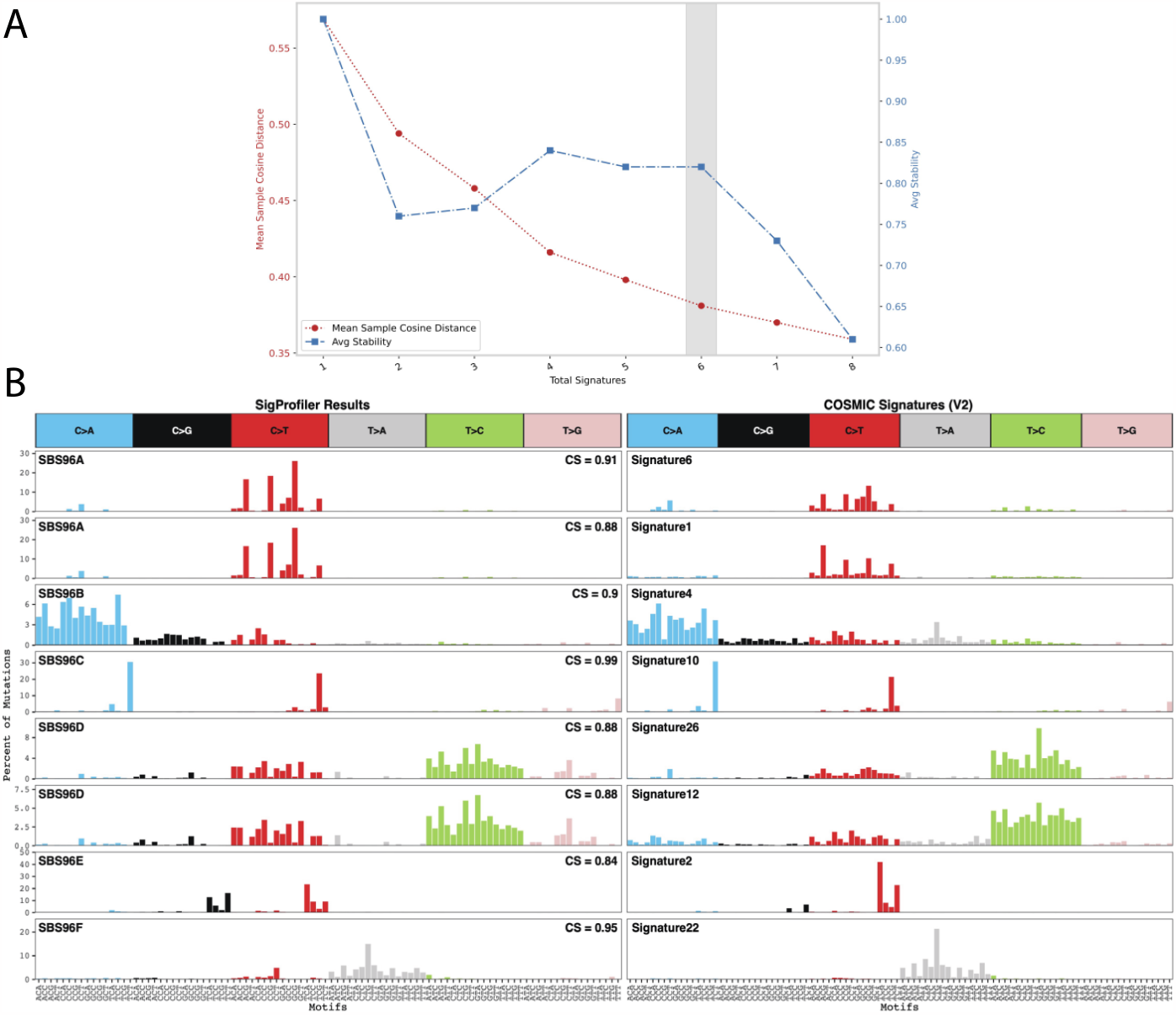
Discovery of mutational signatures *de novo* using NMF. **(A)** NMF in the SigProfiler package was run to identify mutational signatures. The optimal number of signatures was determined to be six based on the maximal difference between the mean sample cosine distance and average stability metrics. **(B)** All discovered signatures were highly correlated with at least one known signature in the COSMIC database showing that no new highly active signatures could be identified in this cohort.

**Supplementary Figure 2.**
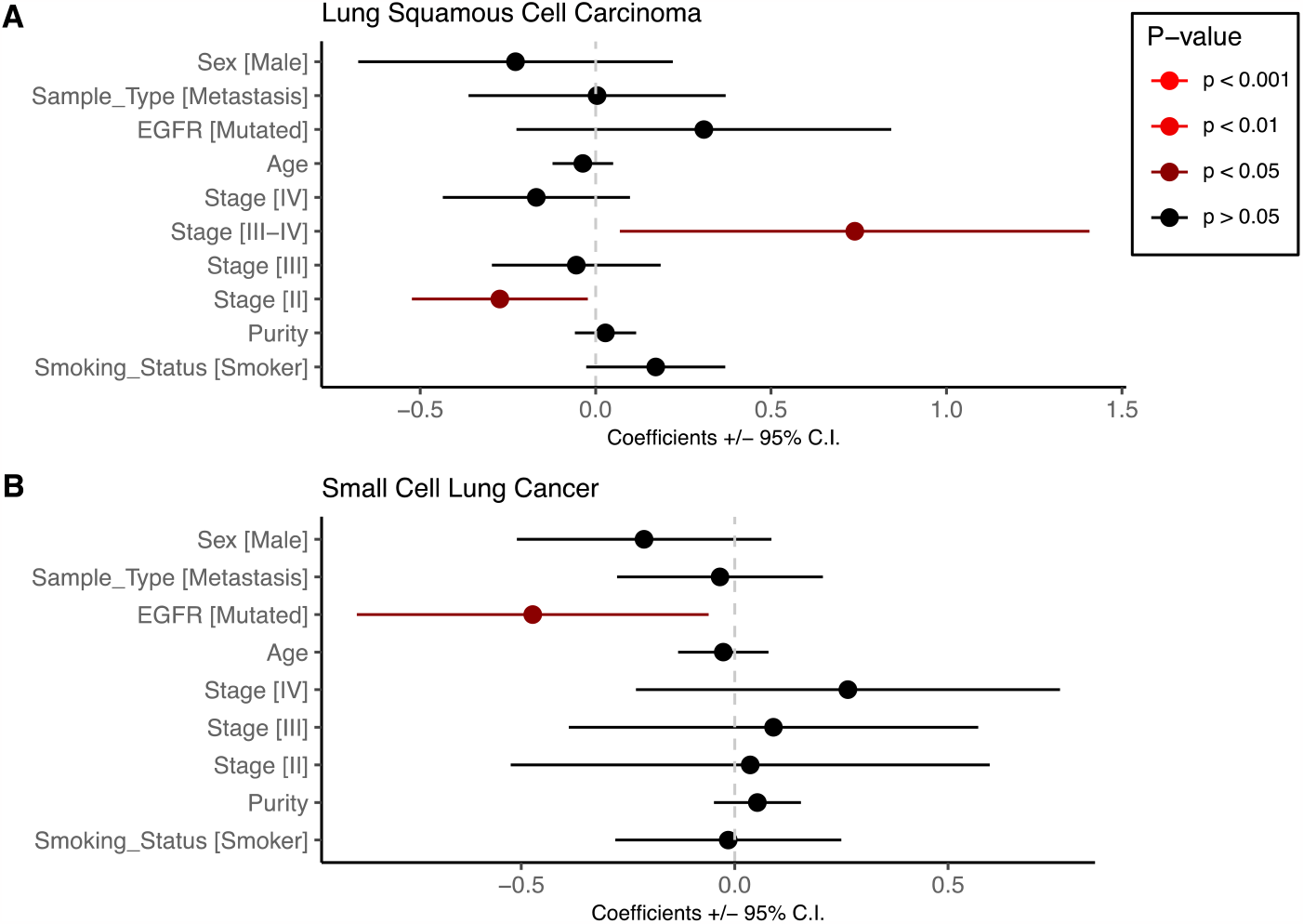
Lack of associations between clinical variables and SBS4 activity in lung squamous cell carcinoma (LUSC) and small cell lung cancer (SCLC). A multivariate linear model was used to assess the relationship between SBS4 activity and clinical variables in **(A)** lung squamous cell carcinoma and **(B)** small cell lung cancer. Only moderate associations were observed between SBS4 activity and Stage II or Stage III-IV tumors in LUSC or *EGFR* mutations in SCLC (p < 0.05). No associations were observed between smoking status and sex (p > 0.05).

